# Antipsychotic management in general practice: serial cross-sectional study (2011-2020)

**DOI:** 10.1101/2024.06.18.24309122

**Authors:** Alan Woodall, Alex Gampel, Huw Collins, Lauren E Walker, Frances S Mair, Sally Sheard, Pyers Symon, Iain Buchan

**Author notes:** Deceased (Huw Collins).

## Abstract

**Background:** Long-term use of antipsychotics confers increased risk of cardiometabolic disease. Ongoing need should be reviewed regularly by psychiatrists.

**Aim:** To explore trends in antipsychotic management in general practice, and proportions of patients prescribed antipsychotics receiving psychiatrist review.

**Design and Setting:** A serial cross-sectional study using linked general practice and hospital data in Wales (2011-2020).

**Method:** Participants were adults (≥18 years) registered with general practices in Wales. Outcome measures were prevalence of patients receiving ≥6 antipsychotic prescriptions annually, proportion of patients prescribed antipsychotics receiving annual psychiatrist review, and proportion of patients prescribed antipsychotics registered on UK Serious Mental Illness, Depression and/or Dementia registers, or not on any of these registers.

**Results:** Prevalence of adults prescribed long-term antipsychotics increased from 1.06% (95%CI 1.04 to 1.07%) in 2011 to 1.45% (95%CI 1.43 to 1.46%) in 2020. The proportion receiving annual psychiatrist review decreased from 59.6% (95%CI 58.9 to 60.4%) in 2011 to 52.0% (95C%CI 51.4 to 52.7%) in 2020. The proportion of antipsychotics prescribed to patients not on the Serious Mental Illness register increased from 50% (95%CI 49 to 51%) in 2011 to 56% (95%CI 56 to 57%) by 2020.

**Conclusions:** Prevalence of long-term antipsychotic use is increasing. More patients are managed by general practitioners without psychiatrist review and are not on monitored disease registers; they thus may be less likely to undergo cardiometabolic monitoring and miss opportunities to optimise or deprescribe antipsychotics. These trends pose risks for patients and need to be addressed urgently.

**How this fits in:** Antipsychotics are effective treatments for serious mental illness, but long-term use increases the risk of cardiometabolic disease, therefore ongoing use should be reviewed regularly. There is increasing use of these medications for non-psychotic mental illness by psychiatrists who then discharge patients to sole care by GPs, often for patients who are not captured on disease registers that automate recall for cardiometabolic risk monitoring. This study demonstrates a rising burden of long-term antipsychotic prescribing in general practice, with nearly half of all adult patients not under annual psychiatrist review, and only 44% of long-term antipsychotic prescriptions being issued to patients coded on the Quality and Outcomes Framework (QOF) Serious Mental Illness register. This means many patients taking antipsychotics may be missed for cardiometabolic screening and expert review. Policy changes are required to ensure regular effective review and monitoring to prevent increasing morbidity and early mortality for patients prescribed long-term antipsychotics.

## Introduction

Antipsychotics are licenced in the United Kingdom (UK) for the management of serious mental illness (SMI) such as schizophrenia and bipolar disorder [1]. Some antipsychotics have UK licences for treatment of depression (quetiapine) and behavioural management in dementia (risperidone) [2]. Antipsychotics are also often prescribed, off-licence and long-term, for personality disorder [3], behavioural management in learning disability [4], autism[5] and anxiety [6], in contravention of guidelines issued by the National Institute of Health and Care Excellence (NICE) [3, 7, 8].

Antipsychotics are effective medications for SMI, but long-term use increases risk of obesity [9], diabetes mellitus [10] and cardiovascular disease [11, 12]. Psychiatrist-led review of antipsychotics should occur regularly to prevent overuse; patients with SMI die 15-20 years prematurely, mainly due to cardiovascular disease and cancer [13–17], but their mortality is higher in many other conditions such as severe COVID-19 infection [18]. Avoidable early mortality for patients with SMI is of significant international public health concern [19]; patients with SMI experience lower screening rates, less monitoring for cardiometabolic risk, and fewer interventions when cardiovascular disease occurs [20]. Poorer physical health outcomes for patients with SMI remain problematic, even in countries with universal healthcare [21]

General practitioners (GPs) in the UK provide physical health reviews of patients with SMI [13, 22]. These are conducted via annual recall of patients with diagnoses that encode them on the Quality Outcomes Framework (QOF) serious mental illness (QOF-SMI) register (termed a ‘mental health’ register, but only includes patients with psychotic illnesses and/or prescribed lithium). QOF is a performance management programme to establish registers of patients with chronic illnesses who require enhanced care, and forms part of the UK GP contract (now excluding Scotland) [23] (Supplementary Table 1). The QOF-SMI annual review includes monitoring of alcohol consumption, smoking, body mass index, blood pressure, blood glucose and lipids, to detect patients at risk of developing cardiometabolic diseases [13]. However, patients taking antipsychotics for dementia, depression, or conditions such as anxiety or personality disorder are not captured on the QOF-SMI register and may not receive cardiometabolic monitoring; a cohort study of 47,724 patients in 2016 found <50% patients prescribed antipsychotics had an SMI diagnosis [7]. GPs report they lack competencies to optimise antipsychotics (e.g., alter dose, switch, or stop) without input from psychiatrists undertaking regular reviews [22, 24, 25]. Patients who are commenced on antipsychotics by psychiatrists are frequently discharged to general practice; 31% of patients with SMI were managed solely by general practice in 2009 [26], and may take antipsychotics long-term without further psychiatric review. We addressed 3 main research questions:

1. Has the prevalence of long-term antipsychotic use in general practice changed over the decade just before the COVID-19 pandemic?
2. Has the proportion of patients prescribed long-term antipsychotics receiving psychiatrist review changed?
3. Has the proportion of people prescribed antipsychotics that are included on QOF- SMI register changed?

## Methods

### Study design

#### A serial cross-sectional population study was undertaken with reference to the

‘Strengthening the Reporting of Observational Studies in Epidemiology’ (STROBE) guidelines [27]. This methodology has been used in international studies of antipsychotic prescribing [28, 29].

### Study population and setting

Individuals registered with GP practices providing data to the Secure Anonymised Information Linkage (SAIL) databank formed the population. Analysis focused on adult use (≥18 years), subdivided by subspeciality: general adult psychiatry (18-64 years) and older adult psychiatry (≥65 years). Patient age was calculated on 1^st^ January each year.

We defined psychiatrist review as occurring where a record of psychiatrist outpatient clinic contacts with a registered medical practitioner or advanced nurse practitioner (including general adult-, older adult-, child and adolescent-, forensic- or learning disability psychiatry, and psychotherapy subspecialties), or psychiatric hospital admission, occurred in the 12 months preceding each 1^st^ January. If neither contact occurred, then we defined antipsychotic management as being solely provided by GPs. Two health boards of the seven in Wales could not provide psychiatric outpatient clinic returns for the study period; therefore, analysis of the proportions of patients prescribed antipsychotics who had received psychiatrist review was limited to five health boards providing clinic returns for 2011-2020. These seven health boards all had similar prevalences of SMI, depression and dementia from QOF returns to Welsh government [30]. The COVID-19 pandemic disrupted normal outpatient services; hence we focused on psychiatrist review up to 1/1/2020.

### Data sources

The SAIL databank was the data source. This is an expanding data repository of 500 million anonymised and encrypted individual-level records from primary and secondary health care sources, and sociodemographic data relevant to health. This includes national datasets covering the whole of Wales (approximately 3 million population) [31–35]. Patients registered with general practices providing data to SAIL were examined using the Welsh Longitudinal General Practice (WLGP) dataset. As of 2023, 87% of Welsh general practices provide historical electronic health record data to SAIL for each patient, covering 83% of the population [36]. Outpatient clinic attendance was determined from the Outpatient Dataset for Wales (OPDW). Psychiatric hospital admission was ascertained using the Patient Episode Dataset for Wales (PEDW). SAIL databank has been validated for use in studies on mental illness [15].

Diagnoses for inclusion in QOF-SMI, depression and dementia registers recorded in GP records and prescriptions for antipsychotics (with a UK licence for use during the study period as listed in the British National Formulary [2]) were extracted using 5-digit Read Codes (v2) from WLGP (see repository https://github.com/alanwoodall/AMP-Epidemiology).

Discussion with two psychiatrists confirmed that two antipsychotics were rarely prescribed for psychiatric purposes and are mainly used for physical health: prochlorperazine (antiemetic) and levomepromazine (palliative care); these were excluded from analysis.

Parenteral (‘depot’) antipsychotics or clozapine issued by GPs were also captured, but these are usually prescribed directly by psychiatrists.

### Analysis

The primary outcome measure was prevalence of adults issued antipsychotics on ≥6 separate days in each calendar year. Analysis by gender, ethnicity and index of deprivation quintile (2019 Welsh Index of Deprivation, WIMD) was also undertaken. We undertook sensitivity analysis of antipsychotic script events per annum to examine patterns of use (see repository); we defined ≥6 script events in a 12 month period as long-term antipsychotic use conservatively, based on the frequency of psychiatric outpatient review (usually biannually) and on definition of regular use in other studies [37]. Further analysis was undertaken to examine antipsychotic by age (18-64 years and ≥65 years), to mirror the UK division between general adult- and old-age psychiatry subspecialities. The prevalence of the six most common antipsychotics prescribed was also determined.

For the proportion of people on antipsychotics receiving psychiatrist review, we undertook sensitivity analysis, varying the period to capture review occurrence up to 5 years. We chose 12 months as the period to define antipsychotic review by psychiatry.

The number of patients who had a lifetime history of diagnostic codes in the QOF SMI, QOF dementia and/or QOF depression registers was determined, along with the number of patients on each disease register prescribed long-term antipsychotics. These three psychiatric illness QOF registers cover diagnoses for almost all licensed uses of antipsychotics in the UK [2]. The number of patients prescribed antipsychotics not on any of these three psychiatric QOF-registers was also determined.

### Statistical analysis

Statistical analysis was undertaken using StatsDirect (Version 4.0.1, www.statsdirect.com). The Clopper-Pearson method was used for confidence intervals of binomial proportions [38]. Differences between binomial proportions were evaluated using the Miettinen-Nurminen method [39]. Results are presented as the main effect with a 95% confidence interval. A 5% significance level was used for hypothesis tests.

### Patient and public involvement

Public advisors for research with the Mental Health Research for Innovation Centre, University of Liverpool, provided oversight of the study.

## Results

### Changes in prevalence of long-term antipsychotic prescribing

Table 1 shows the prevalence of long-term antipsychotic exposure in 2011 and 2020 (also see Supplementary Figure 1). Prevalence of adults ≥18 years exposed to antipsychotics increased from 1.06% to 1.45%; this increase is exclusively in those 18-64 years (0.92% in 2011 increasing to 1.46% in 2020). Of individual antipsychotics, quetiapine was the most prescribed in those ≥18 years (increasing from 0.31% in 2011 to 0.56% in 2020). For those ≥65 years, antipsychotic prevalence fell from 1.51% in 2011 to 1.42% in 2020. Risperidone was the most prescribed antipsychotic in this group (increasing from 0.17% in 2011 to 0.36% in 2020).

**Table 1:**
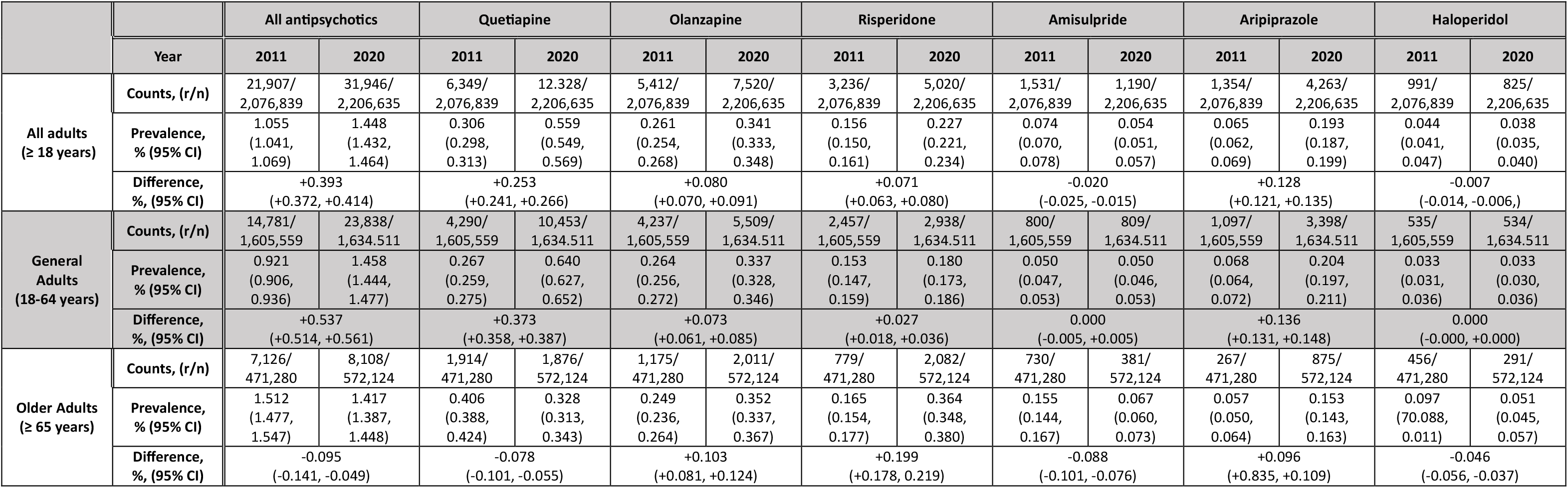
Antipsychotic prevalence changes between 2011 and 2020. Analysis based on number of patients prescribed regular antipsychotics, (r), as a proportion of the at-risk population in a calendar year, (n)

|  | Year | All antipsychotics |  | Quetiapine |  | Olanzapine |  | Risperidone |  | Amisulpride |  | Aripiprazole |  | Haloperidol |  |
| --- | --- | --- | --- | --- | --- | --- | --- | --- | --- | --- | --- | --- | --- | --- | --- |
|  |  | 2011 | 2020 | 2011 | 2020 | 2011 | 2020 | 2011 | 2020 | 2011 | 2020 | 2011 | 2020 | 2011 | 2020 |
| All adults<br>(≥ 18 years) | Counts, (r/n) | 21,907/<br>2,076,839 | 31,946/<br>2,206,635 | 6,349/<br>2,076,839 | 12,328/<br>2,206,635 | 5,412/<br>2,076,839 | 7,520/<br>2,206,635 | 3,236/<br>2,076,839 | 5,020/<br>2,206,635 | 1,531/<br>2,076,839 | 1,190/<br>2,206,635 | 1,354/<br>2,076,839 | 4,263/<br>2,206,635 | 991/<br>2,076,839 | 825/<br>2,206,635 |
|  | Prevalence,<br>% (95% CI) | 1.055<br>(1.041,<br>1.069) | 1.448<br>(1.432,<br>1.464) | 0.306<br>(0.298,<br>0.313) | 0.559<br>(0.549,<br>0.569) | 0.261<br>(0.254,<br>0.268) | 0.341<br>(0.333,<br>0.348) | 0.156<br>(0.150,<br>0.161) | 0.227<br>(0.221,<br>0.234) | 0.074<br>(0.070,<br>0.078) | 0.054<br>(0.051,<br>0.057) | 0.065<br>(0.062,<br>0.069) | 0.193<br>(0.187,<br>0.199) | 0.044<br>(0.041,<br>0.047) | 0.038<br>(0.035,<br>0.040) |
|  | Difference,<br>%, (95% CI) | +0.393<br>(+0.372, +0.414) |  | +0.253<br>(+0.241, +0.266) |  | +0.080<br>(+0.070, +0.091) |  | +0.071<br>(+0.063, +0.080) |  | -0.020<br>(-0.025, -0.015) |  | +0.128<br>(+0.121, +0.135) |  | -0.007<br>(-0.014, -0.006,) |  |
| General<br>Adults<br>(18-64 years) | Counts, (r/n) | 14,781/<br>1,605,559 | 23,838/<br>1,634,511 | 4,290/<br>1,605,559 | 10,453/<br>1,634,511 | 4,237/<br>1,605,559 | 5,509/<br>1,634,511 | 2,457/<br>1,605,559 | 2,938/<br>1,634,511 | 800/<br>1,605,559 | 809/<br>1,634,511 | 1,097/<br>1,605,559 | 3,398/<br>1,634,511 | 535/<br>1,605,559 | 534/<br>1,634,511 |
|  | Prevalence,<br>% (95% CI) | 0.921<br>(0.906,<br>0.936) | 1.458<br>(1.444,<br>1.477) | 0.267<br>(0.259,<br>0.275) | 0.640<br>(0.627,<br>0.652) | 0.264<br>(0.256,<br>0.272) | 0.337<br>(0.328,<br>0.346) | 0.153<br>(0.147,<br>0.159) | 0.180<br>(0.173,<br>0.186) | 0.050<br>(0.047,<br>0.053) | 0.050<br>(0.046,<br>0.053) | 0.068<br>(0.064,<br>0.072) | 0.204<br>(0.197,<br>0.211) | 0.033<br>(0.031,<br>0.036) | 0.033<br>(0.030,<br>0.036) |
|  | Difference,<br>%, (95% CI) | +0.537<br>(+0.514, +0.561) |  | +0.373<br>(+0.358, +0.387) |  | +0.073<br>(+0.061, +0.085) |  | +0.027<br>(+0.018, +0.036) |  | 0.000<br>(-0.005, +0.005) |  | +0.136<br>(+0.131, +0.148) |  | 0.000<br>(-0.000, +0.000) |  |
| Older Adults<br>(≥ 65 years) | Counts, (r/n) | 7,126/<br>471,280 | 8,108/<br>572,124 | 1,914/<br>471,280 | 1,876/<br>572,124 | 1,175/<br>471,280 | 2,011/<br>572,124 | 779/<br>471,280 | 2,082/<br>572,124 | 730/<br>471,280 | 381/<br>572,124 | 267/<br>471,280 | 875/<br>572,124 | 456/<br>471,280 | 291/<br>572,124 |
|  | Prevalence,<br>% (95% CI) | 1.512<br>(1.477,<br>1.547) | 1.417<br>(1.387,<br>1.448) | 0.406<br>(0.388,<br>0.424) | 0.328<br>(0.313,<br>0.343) | 0.249<br>(0.236,<br>0.264) | 0.352<br>(0.337,<br>0.367) | 0.165<br>(0.154,<br>0.177) | 0.364<br>(0.348,<br>0.380) | 0.155<br>(0.144,<br>0.167) | 0.067<br>(0.060,<br>0.073) | 0.057<br>(0.050,<br>0.064) | 0.153<br>(0.143,<br>0.163) | 0.097<br>(0.088,<br>0.111) | 0.051<br>(0.045,<br>0.057) |
|  | Difference,<br>%, (95% CI) | -0.095<br>(-0.141, -0.049) |  | -0.078<br>(-0.101, -0.055) |  | +0.103<br>(+0.081, +0.124) |  | +0.199<br>(+0.178, 0.219) |  | -0.088<br>(-0.101, -0.076) |  | +0.096<br>(+0.835, +0.109) |  | -0.046<br>(-0.056, -0.037) |  |

Table 2 shows prevalence of antipsychotics analysed by sex, deprivation quintile and ethnicity. Females were more likely than males to be prescribed long-term antipsychotics (2020 female/male prevalence ratio 1.10 (95% CI: 1.07-1.12). There was increasing prevalence of antipsychotic use in the most deprived compared to least deprived quintiles (prevalence ratio in 2011 was 2.30, increasing to 2.58 in 2020). When analysed by ethnicity, white individuals had a higher prevalence of antipsychotic use than other ethnic groups, with Asian individuals having the lowest prevalence (2020 prevalence ratio Asian/White: 0.37 (95%CI: 0.33 - 0.42)

**Table 2:** Prevalence of antipsychotic use in 2011 and 2020 by subgroup characteristic of sex, ethnicity and quintile of 2019 Welsh Index of Multiple Deprivation (WIMD) score. Analysis based on number of patients prescribed regular antipsychotics (r), as a proportion of the at-risk population (n), in the calendar year. ^‡^Secondary suppression was used for <10 missing gender codes by imputing the values equally between male and females to reduce risk of disclosure.

|  |  | 2011 |  |  | 2020 |  |  | 2020 - 2011 |
| --- | --- | --- | --- | --- | --- | --- | --- | --- |
| Subgroup |  | Counts<br>(r/n) | Prevalence, %<br>(95%CI) | Prevalence ratio<br>(95%CI) | Counts<br>(r/n) | Prevalence, %<br>(95%CI) | Prevalence ratio<br>(95%CI) | Prevalence difference, %<br>(95% CI) |
| WIMD quintile | 1 (most deprived) | 6,112 /<br>397,630 | 1.54<br>(1.50 – 1.58) | 2.30<br>(2.20 – 2.40) | 9,156 /<br>421,284 | 2.17<br>(2.13 – 2.21) | 2.58<br>(2.48 – 2.68) | 0.64<br>(0.58 – 0.69) |
|  | 2 | 5,136 /<br>411,764 | 1.25<br>(1.22 – 1.28) | 1.87<br>(1.78 – 1.95) | 7,033 /<br>419,569 | 1.68<br>(1.64 – 1.72) | 1.99<br>(1.91 – 2.07) | 0.43<br>(0.38 – 0.48) |
|  | 3 | 3,828 /<br>407,578 | 0.94<br>(0.91 – 0.97) | 1.40<br>(1.33 – 1.46) | 5,069 /<br>410,416 | 1.24<br>(1.21 – 1.27) | 1.46<br>(1.40 – 1.53) | 0.30<br>(0.25 – 0.34) |
|  | 4 | 3,200 /<br>377,925 | 0.85<br>(0.82 – 0.88) | 1.27<br>(1.21 – 1.33) | 4,284 /<br>390,523 | 1.10<br>(1.07 – 1.13) | 1.30<br>(1.24 – 1.36) | 0.25<br>(0.21 – 0.29) |
|  | 5 (least deprived) | 2,843 /<br>425,334 | 0.67<br>(0.65 – 0.69) | 1.00<br>(0.95 – 1.05) | 3,582 /<br>424,598 | 0.84<br>(0.81 – 0.87) | 1.00<br>(0.96 – 1.05) | 0.18<br>(0.14 – 0.21) |
|  | Missing | 788 /<br>56,608 | 1.39<br>(1.30 – 1.49) | 2.08<br>(1.93 – 2.25) | 2,822 /<br>140,245 | 2.01<br>(1.94 – 2.08) | 2.39<br>(2.27 – 2.51) | 0.62<br>(0.50 – 0.74) |
| Gender<sup>†</sup> | Female | 11,741 /<br>1,048,240 | 1.12<br>(1.10 – 1.14) | 1.13<br>(1.10 – 1.16) | 16,826 /<br>1,114,501 | 1.51<br>(1.49 – 1.53) | 1.09<br>(1.07 – 1.12) | 0.39<br>(0.36 – 0.42) |
|  | Male | 10,166 /<br>1,028,599 | 0.99<br>(0.97 – 1.01) | 1.00<br>(0.97 – 1.03) | 15,120 /<br>1,092,134 | 1.38<br>(1.36 – 1.40) | 1.00<br>(0.98 – 1.02) | 0.40<br>(0.37 – 0.43) |
| Ethnicity | Asian | 142 /<br>29,749 | 0.48<br>(0.41 – 0.57) | 0.33<br>(0.28 – 0.39) | 273 /<br>37,339 | 0.73<br>(0.65 – 0.82) | 0.37<br>(0.33 – 0.42) | 0.25<br>(0.14 – 0.37) |
|  | Black | 118 /<br>9,854 | 1.20<br>(1.00 – 1.43) | 0.83<br>(0.69 – 0.99) | 185 /<br>12,876 | 1.44<br>(1.25 – 1.66) | 0.73<br>(0.63 – 0.84) | 0.24<br>(-0.06 – 0.05) |
|  | Mixed | 65 /<br>6,102 | 1.07<br>(0.84 – 1.36) | 0.74<br>(0.58 – 0.94) | 126 /<br>8,712 | 1.45<br>(1.22 – 1.73) | 0.73<br>(0.62 – 0.87) | 0.38<br>(0.01 – 0.74) |
|  | Other | 60 /<br>8,652 | 0.69<br>(0.53 – 0.89) | 0.48<br>(0.37 – 0.62) | 133 /<br>13,635 | 0.98<br>(0.83 – 1.16) | 0.50<br>(0.42 – 0.59) | 0.28<br>(0.03 – 0.52) |
|  | White | 17,309 /<br>1,200,276 | 1.44<br>(1.42 – 1.46) | 1.00<br>(0.98 – 1.02) | 24,862 /<br>1,261,529 | 1.97<br>(1.95 – 1.99) | 1.00<br>(0.98 – 1.02) | 0.53<br>(0.50 – 0.56) |
|  | Missing | 4,213 /<br>822,176 | 0.51<br>(0.49 – 0.53) | 0.36<br>(0.34 – 0.37) | 6,367 /<br>872,544 | 0.73<br>(0.71 – 0.75) | 0.37<br>(0.36 – 0.38) | 0.22<br>(0.19 – 0.24) |

Supplementary Table 2 shows the counts of patients receiving long-term antipsychotics listed in the British National Formulary in 2011 and 2020, and their licensed indications. The proportion of long-term antipsychotic exposure that is accounted for by ‘second generation’ or ‘atypical’ antipsychotics increased from 79.7% in 2011 to 91.7% in 2020. Approximately 94% of antipsychotic exposures in 2020 are accounted for by 6 individual antipsychotics: quetiapine (37.3%), olanzapine (22.6%), risperidone (15.3%) aripiprazole (12.9%), amisulpride (3.6%) and haloperidol (2.5%).

### Proportion of patients prescribed long-term antipsychotics receiving psychiatric specialist review

The percentage of adults ≥18 years taking long-term antipsychotics who received psychiatrist review in the 12 months preceding each year (2011-2020) fell from 59.6% to 52.0% (Table 3; also see Supplementary Figure 2). The proportion of 18–64-year-olds taking long-term antipsychotics that received psychiatrist review decreased from 65.7% in 2011 to 55.0% in 2020. The greatest decrease in psychiatrist review occurred in patients prescribed quetiapine (-17.0%). For patients ≥65 years, less than half underwent psychiatrist review, decreasing from 47.5% in 2011 to 43.4% in 2020. The greatest decrease was seen for those taking amisulpride (-18.2%).

**Table 3:** Proportion of patients taking long-term antipsychotics who have had a specialist psychiatrist review in previous 12 months. Analysis based on number of patients prescribed antipsychotics who had received psychiatrist review, (r), as a proportion of the number of patients prescribed antipsychotics in that calendar year, (n).

|  | Year | All antipsychotics |  | Quetiapine |  | Olanzapine |  | Risperidone |  | Amisulpride |  | Aripiprazole |  | Haloperidol |  |
| --- | --- | --- | --- | --- | --- | --- | --- | --- | --- | --- | --- | --- | --- | --- | --- |
|  |  | 2011 | 2020 | 2011 | 2020 | 2011 | 2020 | 2011 | 2020 | 2011 | 2020 | 2011 | 2020 | 2011 | 2020 |
| Adults<br>(≥18 years) | Patients reviewed by psychiatrist, r/n | 9,574/<br>16,053 | 11,894/<br>22,855 | 2,978/<br>4,786 | 4,539/<br>8,940 | 2,362/<br>3,698 | 2,736/<br>5,272 | 1,428/<br>2,466 | 1,750/<br>3,607 | 751/<br>1,146 | 504/<br>919 | 787/<br>974 | 2,102/<br>3,191 | 390/<br>813 | 311/<br>593 |
|  | Patients reviewed by psychiatrist, %, (95% CI) | 59.6<br>(58.9-60.4) | 52.0<br>(51.4-52.7) | 62.2<br>(60.8-63.6) | 50.8<br>(49.7-51.8) | 63.9<br>(62.3-65.4) | 51.9<br>(50.6-53.2) | 57.9<br>(56.0-59.8) | 48.5<br>(46.9-50.2) | 65.5<br>(62.7-68.2) | 54.8<br>(51.6-58.0) | 80.8<br>(78.2-83.2) | 65.9<br>(64.2-67.5) | 48.0<br>(44.6-51.4) | 52.5<br>(48.4-56.5) |
|  | Difference, %, (95% CI) | -7.6 (-8.6, -6.6) |  | -11.4 (-13.2, -9.7) |  | -12.0 (-14.0, -9.9) |  | -9.4 (-12.0, -6.8) |  | -10.7 (-14.9, -6.5) |  | -14.9(-17.8, -11.9) |  | +4.5 (-0.8, +9.8) |  |
| General Adults<br>(18-64 years) | Patients reviewed by psychiatrist, r/n | 7,028/<br>10,691 | 9,358/<br>17,013 | 2,315/<br>3,339 | 3,945/<br>7,538 | 1,901/<br>2,900 | 2,104/<br>3,882 | 1,124/<br>1,903 | 1,110/<br>2,141 | 432/<br>585 | 384/<br>609 | 679/<br>818 | 1,775/<br>2,593 | 263/<br>431 | 223/<br>357 |
|  | Patients reviewed by psychiatrist, %, (95% CI) | 65.7<br>(64.8-66.6) | 55.0<br>(54.3-55.8) | 69.3<br>(67.8-70.9) | 52.3<br>(51.2-53.5) | 65.6<br>(63.8-67.3) | 54.2<br>(52.6-55.8) | 59.1<br>(56.9-61.3) | 51.8<br>(49.7-54.0) | 73.8<br>(70.3-77.4) | 63.1<br>(59.2-66.9) | 83.0<br>(80.4-85.6) | 68.5<br>(66.7-70.2) | 61.0<br>(56.4-65.6) | 62.5<br>(57.4-67.5) |
|  | Difference, %, (95% CI) | -10.7 (-11.7, -9.5) |  | -17.0 (-19.0, -15.0) |  | -11.4 (-13.7, -9.0) |  | -7.3 (-10.3, -4.2) |  | -10.7 (-16.1, -5.5) |  | -14.5 (-18.1, -11.0) |  | +1.5 (-8.3, +5.4) |  |
|  | Patients reviewed by psychiatrist, r/n | 2,546/<br>5,362 | 2,536/<br>5,842 | 663/<br>1,447 | 594/<br>1,402 | 461/<br>798 | 632/<br>1,370 | 304/<br>563 | 640/<br>1,466 | 319/<br>561 | 120/<br>310 | 108/<br>156 | 327/<br>598 | 127/<br>382 | 88/<br>236 |
| Older Adults<br>(≥65 years) | Patients reviewed by psychiatrist, %, (95% CI) | 47.5<br>(46.1-48.8) | 43.4<br>(42.1-44.7) | 45.8<br>(43.3-48.4) | 42.4<br>(39.8-45.0) | 57.8<br>(54.3-61.2) | 46.1<br>(43.5-48.8) | 54.0<br>(49.9-58.1) | 43.7<br>(41.1-46.2) | 56.9<br>(52.8-61.0) | 38.7<br>(33.3-44.1) | 69.2<br>(62.0-76.5) | 54.7<br>(50.7-58.7) | 33.2<br>(28.5-38.0) | 37.3<br>(31.1-43.5) |
|  | Difference, %, (95% CI) | -4.1 (-5.9, -2.2) |  | -3.4 (-7.1, +2.0) |  | -11.7 (-16.0, -7.3) |  | -10.3 (-15.2, -5.5) |  | -18.2 (-25.1, -11.2) |  | -14.5 (-23.3, -5.8) |  | +4.1 (-11.8, +3.7) |  |

We conducted sensitivity analysis of psychiatrist review by varying the period of time allowed up to 5 years prior to 1/1/2020 (Supplementary Figure 3). For patients aged 18-64 years prescribed long-term antipsychotics in 2020, 22.9% had no psychiatrist contact in the previous 5 years. For patients aged >65 years, 29.9% of those taking antipsychotics long-term in 2020 had no psychiatrist contact in the previous 5 years.

### Proportions of patients on the psychiatric QOF registers prescribed long-term antipsychotics

In 2020, 1 in 63 (1.59%) adults ≥18 years had a lifetime diagnostic code for inclusion on the QOF SMI register, (+0.17% since 2011); 1 in 5 adults (20.7%) had a code for inclusion on the QOF depression register (+5.6% since 2011); 1 in 130 adults (0.77%) had a code for inclusion on the QOF dementia register (+ 0.1% since 2011) (Table 4). For adults without codes for inclusion on these three psychiatric illness QOF registers, the proportion fell from 83.7% in 2011, to 78.1% in 2020

**Table 4:**
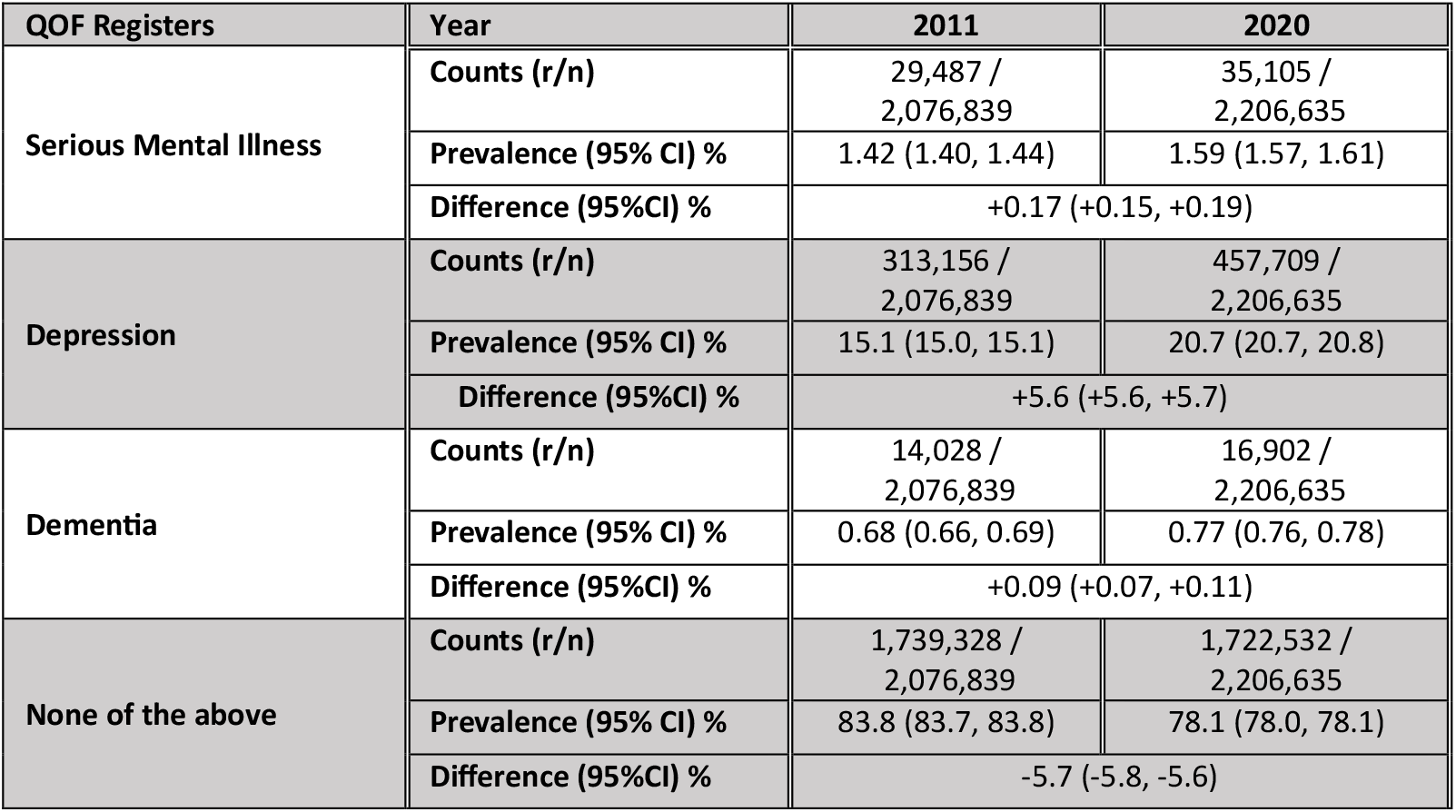
Quality and Outcomes Framework registration prevalence changes for persons aged ≥18 years between 2011 and 2020. . Analysis is based on the number of patients, (r), who have ever been the relevant register, as a proportion of the at-risk population, (n), in the calendar year.

| QOF Registers | Year | 2011 | 2020 |
| --- | --- | --- | --- |
| Serious Mental Illness | Counts (r/n) | 29,487 /<br>2,076,839 | 35,105 /<br>2,206,635 |
|  | Prevalence (95% CI) % | 1.42 (1.40, 1.44) | 1.59 (1.57, 1.61) |
|  | Difference (95%CI) % | +0.17 (+0.15, +0.19) |  |
| Depression | Counts (r/n) | 313,156 /<br>2,076,839 | 457,709 /<br>2,206,635 |
|  | Prevalence (95% CI) % | 15.1 (15.0, 15.1) | 20.7 (20.7, 20.8) |
|  | Difference (95%CI) % | +5.6 (+5.6, +5.7) |  |
| Dementia | Counts (r/n) | 14,028 /<br>2,076,839 | 16,902 /<br>2,206,635 |
|  | Prevalence (95% CI) % | 0.68 (0.66, 0.69) | 0.77 (0.76, 0.78) |
|  | Difference (95%CI) % | +0.09 (+0.07, +0.11) |  |
| None of the above | Counts (r/n) | 1,739,328 /<br>2,076,839 | 1,722,532 /<br>2,206,635 |
|  | Prevalence (95% CI) % | 83.8 (83.7, 83.8) | 78.1 (78.0, 78.1) |
|  | Difference (95%CI) % | -5.7 (-5.8, -5.6) |  |

Table 5 shows the proportion of patients ≥18 years prescribed long-term antipsychotics in each QOF register (also see Supplementary Figure 4). The proportion of patients on the QOF SMI register prescribed antipsychotics increased to 39.6% in 2020 (+2.6% since 2011), with olanzapine the most common exposure in 2020 (11.9%; +0.08% since 2011). The proportion of patients on the QOF dementia register prescribed antipsychotics decreased from 16.7% in 2011 to 12.9% in 2020, with risperidone the most common exposure in this group in 2020 (5.45%; +3.85% since 2011). The proportion of patients on the QOF depression register prescribed antipsychotics increased to 4.12% in 2020 (+0.75% since 2011), with quetiapine the most common exposure in 2020 (1.91%; +0.78% since 2011). The proportion of patients not on any of these three QOF psychiatric illness registers prescribed antipsychotics increased from 0.36% in 2020 (+0.10% since 2011), with quetiapine the most common exposure in 2020 (0.13%, +0.06% since 2011).

**Table 5:**
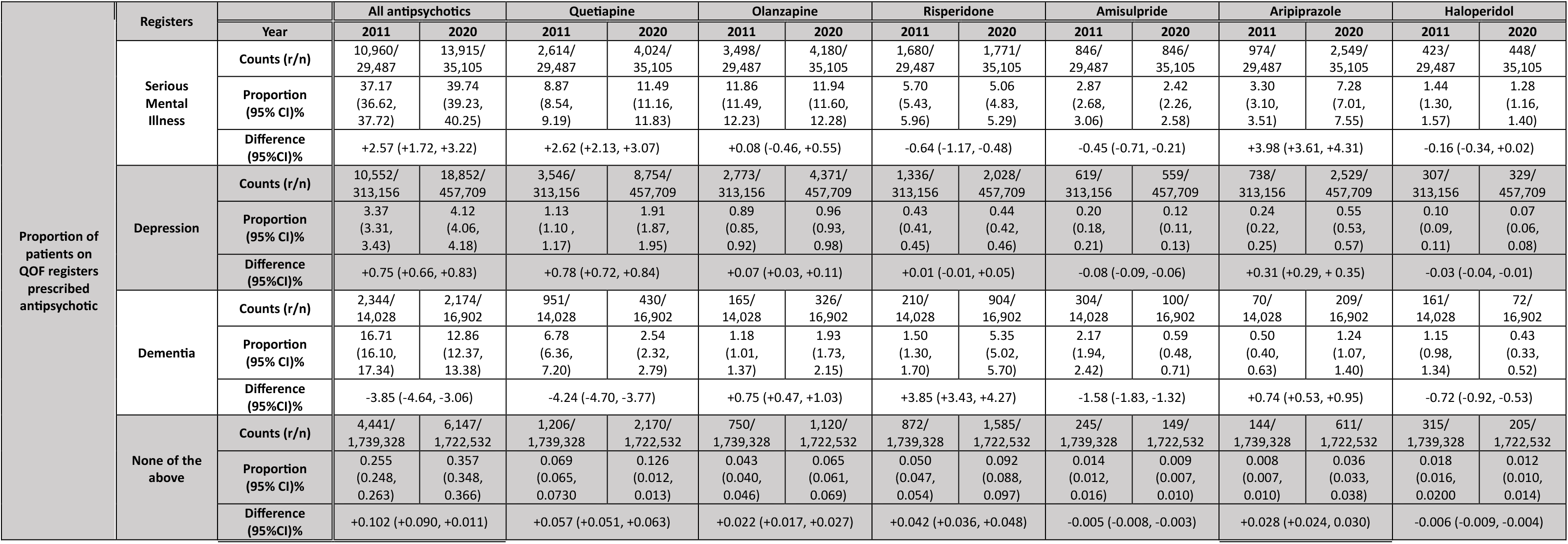
Changes in long-term antipsychotic use in adult patients (≥18 years) with a lifetime code on the QOF Serious Mental Illness, depression and/or dementia registers, and those who are not on any of these three psychiatric illness QOF registers, between 2011 and **2020**. The number of patients with a lifetime code on the relevant QOF registers exposed to antipsychotics, (r), is expressed as a proportion of the total population with a lifetime code for inclusion on each QOF register, (n).

### Proportion of overall long-term antipsychotic use by QOF register status

Table 6 shows the proportion of overall long-term antipsychotic use for patients who are registered on the QOF SMI, QOF dementia, and/or QOF depression registers, and the proportion of overall long-term antipsychotic use for patients not on any of these three QOF registers (also see Supplementary Figure 5). The proportion of overall antipsychotic use for patients on the QOF SMI register fell to 43.6% by 2020 (-7.4% since 2011); the lowest proportion was quetiapine (32.6% in 2020). The proportion of overall antipsychotic use for patients on the QOF dementia register fell to 6.8% in 2020 (- 3.9% since 2011); 18.0% of total risperidone use was for patients on the QOF dementia register. The proportion of antipsychotic use for patients on the QOF depression register increased to 59.0% by 2020 (+10.8% since 2011); 71.0% of total quetiapine use in 2020 was for patients who have ever been on the QOF depression register, a 15.2% increase from 2011 to 2020. The proportion of antipsychotic prescribed for patients not on these three registers fell slightly to 19.2% in 2020 (-1.0% since 2011). The proportion of overall haloperidol use was greatest for this group (34.6% in 2011, decreasing to 24.9% in 2020).

**Table 6:** Changes in proportion of overall antipsychotic use in adult patients (≥18 years) with a lifetime code on the QOF Serious Mental Illness, depression and/or dementia registers, and those who are not on any of these three psychiatric illness QOF registers, between 2011 **and 2020**. The number of patients with a lifetime code on the relevant QOF registers exposed to antipsychotics, (r), is expressed as a proportion of the total number of patients exposed to antipsychotics, (n).

|  | QOF Registers | Year | All antipsychotics |  | Quetiapine |  | Olanzapine |  | Risperidone |  | Amisulpride |  | Aripiprazole |  | Haloperidol |  |
| --- | --- | --- | --- | --- | --- | --- | --- | --- | --- | --- | --- | --- | --- | --- | --- | --- |
|  |  |  | 2011 | 2020 | 2011 | 2020 | 2011 | 2020 | 2011 | 2020 | 2011 | 2020 | 2011 | 2020 | 2011 | 2020 |
| Proportion of total antipsychotic use by QOF register | Serious Mental Illness | Counts (r/n) | 10,960/<br>21,907 | 13,915/<br>31,946 | 2,614/<br>6,349 | 4,024/<br>12,328 | 3,498/<br>5,412 | 4,180/<br>7,520 | 1,680/<br>3,236 | 1,771/<br>5,020 | 846/<br>1,530 | 846/<br>1,190 | 974/<br>1,354 | 2,549/<br>4,263 | 423/<br>911 | 448/<br>825 |
|  |  | Prevalence (95% CI) % | 50.03<br>(49.37, 50.69) | 43.56<br>(43.01, 44.10) | 41.17<br>(39.97, 42.39) | 32.64<br>(31.82, 33.47) | 64.63<br>(63.35, 65.90) | 55.59<br>(54.46, 56.71) | 51.92<br>(50.19, 53.63) | 35.28<br>(33.97, 36.61) | 55.29<br>(52.79, 57.77) | 71.09<br>(68.45, 73.67) | 71.94<br>(69.48, 74.33) | 59.79<br>(58.31, 61.27) | 46.43<br>(43.22, 49.68) | 54.30<br>(50.89, 57.67) |
|  |  | Difference (95%CI) % | -6.47 (-7.33, -5.61) |  | -8.53 (-9.98, -7.08) |  | -9.04 (-10.77, -7.33) |  | -16.64 (-18.82, -14.46) |  | +15.80 (+12.12, +19.47) |  | -12.15 (-15.10, -9.18) |  | +7.87 (+3.16, +12.58) |  |
|  | Depression | Counts (r/n) | 10,552/<br>21,907 | 18,852/<br>31,946 | 3,546/<br>6,349 | 8,754/<br>12,328 | 2,773/<br>5,412 | 4,371/<br>7,520 | 1,336/<br>3,236 | 2,028/<br>5,020 | 619/<br>1,530 | 559/<br>1,190 | 738/<br>1,354 | 2,529/<br>4,263 | 307/<br>911 | 329/<br>825 |
|  |  | Prevalence (95% CI) % | 48.17<br>(47.51, 48.83) | 59.01<br>(58.47, 59.55) | 55.85<br>(54.63, 57.07) | 71.01<br>(70.21, 71.80) | 51.24<br>(49.91, 52.57) | 58.18<br>(57.01, 59.24) | 41.29<br>(39.59, 42.98) | 40.40<br>(39.05, 41.76) | 40.46<br>(38.00, 42.92) | 46.97<br>(44.15, 49.82) | 54.51<br>(51.84, 57.16) | 59.32<br>(57.85, 60.80) | 33.70<br>(30.70, 36.83) | 39.98<br>(36.59, 43.26) |
|  |  | Difference (95%CI) % | +10.84 (+9.99, +11.70) |  | +15.16 (+13.72, +16.59) |  | +6.94 (+5.15, +8.63) |  | -0.89 (-3.06, +1.28) |  | +6.51 (+2.76, +10.27) |  | +4.81 (+1.80, +7.84) |  | +6.28 (+1.64, +10.72) |  |
|  | Dementia | Counts (r/n) | 2,344/<br>21,907 | 2,174/<br>31,946 | 951/<br>6,349 | 430/<br>12,328 | 165/<br>5,412 | 326/<br>7,520 | 210/<br>3,236 | 904/<br>5,020 | 304/<br>1,530 | 100/<br>1,190 | 70/<br>1,354 | 209/<br>4,263 | 161/<br>911 | 72/<br>825 |
|  |  | Prevalence (95% CI) % | 10.70<br>(10.29, 11.11) | 6.81<br>(6.53, 7.09) | 14.98<br>(14.11, 15.88) | 3.49<br>(3.18, 3.83) | 3.05<br>(2.62, 3.54) | 4.34<br>(3.90, 4.82) | 6.49<br>(5.69, 7.39) | 18.01<br>(16.97, 19.07) | 19.87<br>(17.95, 21.94) | 8.40<br>(6.96, 10.13) | 5.17<br>(4.11, 6.48) | 4.90<br>(4.29, 5.59) | 17.67<br>(15.33, 20.28) | 8.73<br>(7.00, 10.85) |
|  |  | Difference (95%CI) % | -3.89 (-4.37, -3.42) |  | -11.49 (-12.28, -10.70) |  | +1.29 (+0.60, +1.94) |  | +11.52 (+10.01, +13.03) |  | -11.47 (-14.16, -8.77) |  | -0.27 (-1.60, +1.06) |  | -8.94 (-12.16, -5.73) |  |
|  | None of the above | Counts (r/n) | 4,441/<br>21,907 | 6,147/<br>31,946 | 1,206/<br>6,349 | 2,170/<br>12,328 | 750/<br>5,412 | 1,120/<br>7,520 | 872/<br>3,236 | 1,585/<br>5,020 | 245/<br>1,530 | 149/<br>1,190 | 144/<br>1,354 | 611/<br>4,263 | 315/<br>911 | 205/<br>825 |
|  |  | Prevalence (95% CI) % | 20.27<br>(19.74, 20.81) | 19.24<br>(18.81, 19.67) | 19.00<br>(18.05, 19.98) | 17.60<br>(16.94, 18.28) | 13.86<br>(12.96, 14.80) | 14.89<br>(14.11, 15.72) | 26.95<br>(25.42, 28.48) | 31.57<br>(30.29, 32.86) | 16.01<br>(14.26, 17.94) | 12.52<br>(10.76, 14.52) | 10.64<br>(9.10, 12.39) | 14.33<br>(13.31, 15.42) | 34.58<br>(31.56, 37.72) | 24.85<br>(22.02, 27.91) |
|  |  | Difference (95%CI) % | -1.03 (-1.71, -0.35) |  | -1.70 (-2.56, -0.23) |  | +1.03 (-0.19, +2.27) |  | +4.62 (+2.61, +6.65) |  | -3.49 (-6.16, -0.83) |  | +3.69 (+1.61, +5.78) |  | -9.73 (-14.05, -5.41) |  |

## Discussion

### Summary

#### This study shows

1. Long-term antipsychotic prevalence increased in adults, with rises in those 18-64 years, and decreased in the ≥65 years population. Prevalence was higher in females, those from the most deprived fifth of the population, and white ethnicity groups.
2. For adults prescribed long-term antipsychotics, the proportion who had annual psychiatrist review decreased from 60% to 52%; this decrease was more pronounced among older patients.
3. The proportion of patients prescribed long-term antipsychotics who are not on the QOF Serious Mental Illness register increased from 50% in 2011 to 56% by 2020, meaning fewer patients would receive QOF recall for cardiometabolic monitoring.

Our work suggests that antipsychotics are increasingly used for unlicensed and/or non-psychotic conditions. The decrease in psychiatrist reviews of patients prescribed antipsychotics, along with increasing use for conditions that are not on the QOF SMI register, may act synergistically to increase risk of people becoming ‘trapped’ on antipsychotics in general practice and increasing their risk of cardiometabolic morbidity. This risk is worse for more deprived patients - antipsychotic prevalence was 2.58 (2.48 - 2.68) times higher in the most deprived compared with the least deprived fifth - and these inequalities have increased over the past decade. Given the prevalence of cardiometabolic disease is highest in the most deprived populations [40], this compounds the multimorbidity risk for this group.

### Strengths and limitations

We believe this is the first study of long-term antipsychotic use to report the proportions of patients who are managed solely in general practice, the first to compare antipsychotic prescribing patterns between different QOF psychiatric illness registers, and of patients not on any psychiatric illness registers. Strengths include: (1) we focus on patients prescribed long-term antipsychotics who are most at risk of cardiometabolic disease; (2) SAIL databank allows patient record linkage between general practice and secondary care datasets, so identifying management trends between general practice and psychiatric services.

Limitations include: (1) SAIL obtains data exclusively from Wales, so caution is needed extrapolating findings to other populations. (2) SAIL cannot extract number of tablets issued per script, so accurate quantification of antipsychotic burden is not possible. (3) SAIL only captures antipsychotic exposure when patients receive prescriptions from NHS general practice; it does not capture prescriptions issued from private services, for patients registered with prison or military general practices, and when scripts are issued by secondary care, as is the case for most parenteral antipsychotic prescriptions. Additionally, clozapine is licensed in the UK only for issue by psychiatrists so would not be routinely captured; only 8 patients received long-term clozapine scripts from general practice in 2020 (Table S2), but the clozapine monitoring service for Wales had 1,984 patients registered in 2019 [41]; these represent missing data of antipsychotic exposure we are not able to capture. (4)We have not identified patients prescribed ≥2 antipsychotics concurrently. (5) For psychiatrist review, we cannot identify ‘indirect’ management between GPs and psychiatrists (e.g. communication without patient contact), nor will it have identified patient contact with non-prescribing mental health professionals, who may provide indirect input into antipsychotic review. (6) While we have used lifetime prevalence of diagnostic codes to determine patient exposure to QOF SMI, QOF Dementia and QOF Depression registers, as other studies have done [7], this will not detect changes in psychiatric diagnoses that can happen over time. Further, psychiatric codes may be inaccurate [42]. Erroneous diagnoses may not be removed from GP records when a subsequent correct one is added [43].

Incomplete, inaccurate or non-contemporaneous coding of psychiatric conditions to measure effectiveness of registers to improve health outcomes is a concern of many UK and international studies, and must be considered when drawing conclusions from data [44–47]. (7) Finally, limited ethnicity coding capture for the population within SAIL databank (61% had ethnicity coding in 2020 using primary and secondary care datasets) reduced our ability to draw conclusions for this subgroup, although the tentative trends we report of similar or higher rates of antipsychotic use for whites compared with other ethnic groups have been previously reported [48]. Studies confirm Wales has relatively poor primary care coding of ethnicity (40%) compared with England (75%), and work is underway to improve ethnicity recording in SAIL to nearly 95% [49].

### Comparison with existing literature

We found an increased prevalence of adults prescribed long-term antipsychotics. The prevalences are consistent with other studies; Marston *et al* reported that 0.7% of the population had exposure to antipsychotics in their cohort study which ended in 2011 [7]; Shoham *et al* reported antipsychotic prevalences of 1.2% in 2014, from a questionnaire survey of patients aged ≥16 years [50]. The greatest increase is for quetiapine, which accounted for 44% of all antipsychotic exposures in the 18-64 years group by 2020. This mirrors findings from international cross-sectional studies: Wilkinson *et al* found prevalence had risen from 1.88% to 2.81% between 2008 and 2015 in New Zealand [28]; Hálfdánarson *et al* report increasing prevalence in 16 countries ranging from 0.32% (Columbia) and 7.8% (Taiwan) by 2014 [29] ; these studies report antipsychotic prescribing from primary and secondary care. Despite this increase in antipsychotic use, psychiatric illness QOF register prevalences have not increased proportionately; this suggests increasing antipsychotic use for non-psychotic disorders, such as anxiety, depression and personality disorder, as has been reported [3, 7, 43]. Limited access to psychological therapies may be increasing antipsychotic use for non-psychotic disorders [22]. Prevalence of long-term antipsychotic exposure in patients ≥65 years has decreased slightly, with significant falls in patients with dementia (-3.85% (95%CI: -4.64 to -3.06%); this is promising, given the increased risk of mortality when antipsychotics are used in Alzheimer’s dementia[51]. However, long-term risperidone prevalence in dementia has increased (+3.85% (95%CI +3.43, +4.27). The UK licence for risperidone is for *’short-term treatment (up to 6 weeks) of persistent aggression in patients with moderate to severe Alzheimer’s dementia‘’* [2], and yet 1 in 19 patients with dementia were prescribed risperidone long-term; studies report pressure on clinicians to use antipsychotics for patients with dementia in care homes[22]. Recent studies suggest this trend of decreasing overall antipsychotic use in patients with dementia was not maintained during the COVID pandemic [52], and international studies have not all reported this decrease [28, 29]. We suspect the UK decrease in long-term antipsychotic prescribing in those ≥65 years we observed was a result of UK governments utilising national prescribing indicators to limit use of antipsychotics in the dementia population to reduce risk of stroke [53].

We found decreasing proportions of patients prescribed antipsychotics have annual psychiatrist review. One explanation may be more antipsychotics are being prescribed by GPs independently; we cannot determine which clinicians initiated antipsychotics, but qualitative studies suggest GPs lack confidence to initiate and manage antipsychotics without psychiatrist input, so we suspect this is unlikely [22, 25, 54]. Another possible explanation is that more patients are discharged on antipsychotics by psychiatrists to general practice, either when patients with SMI are ‘stable’ from a psychiatric perspective, or when antipsychotics are prescribed to patients who do not have psychotic illness.

While the prevalence of antipsychotic exposure is increasing, the proportion of total antipsychotic use for patients with a history of SMI is decreasing; only 44% of antipsychotic use in 2020 was for patients with a history of SMI, similar to the proportion used for SMI (36%) reported by Marston *et al* [7]. This supports our concern that more antipsychotics are being used long-term for non-psychotic disorders in contravention of NICE guidelines (e.g. for personality disorder, where antipsychotics should only be used short-term) [55]. Our data shows the greatest increase in antipsychotic prescribing was for patients with a history of depression; whether antipsychotics are used solely to treat depressive disorders, or the patent has a co-morbid SMI or dementia code cannot be determined; cohort studies are needed to examine antipsychotic use for patients with exclusive depressive diagnoses to determine trends.

Only the QOF SMI register funds GPs to undertake cardiometabolic monitoring of patients taking antipsychotics; 56% of patients prescribed long-term antipsychotics are not on this register, and GPs are not funded to undertake cardiometabolic monitoring for them. We suspect many GPs still perform unfunded cardiometabolic monitoring when antipsychotics are used in non-SMI conditions. However, with most cardiometabolic monitoring devolved to GP practices, even when a patient remains under psychiatrist care, there is a risk that patients taking antipsychotics long-term may not have cardiometabolic monitoring undertaken, as general practice utilises automated QOF SMI register recalls. Patients on the QOF SMI register will get annual recalls for cardiometabolic monitoring and GP review. For patients on the QOF dementia or QOF depression registers taking antipsychotics, there is no QOF requirement to undertake cardiometabolic monitoring, so this is less likely to happen. For patients prescribed long-term antipsychotics not captured on any QOF psychiatric illness register (e.g. personality disorder, anxiety, or autism), there is no funding or automated recall for GP review or cardiometabolic monitoring. GPs are supposed to conduct annual medication reviews, but research shows that these are often of variable effectiveness, particularly when antipsychotics are prescribed and there is no ongoing psychiatric input [22, 25, 54, 56]. A recent study in Scotland shows specialist pharmacist-led physical-health monitoring of patients prescribed antipsychotics can improve cardiometabolic monitoring and proactive intervention to ameliorate risk factors [57].

### Implications for research and/or practice

This study demonstrated a rising burden of antipsychotic use in UK general practice. Further research is required to determine antipsychotic initiation rates by general practitioners as opposed to psychiatrists, and to determine antipsychotic prevalence in different ethnic groups, and specific use of antipsychotics for non-psychotic conditions such as anxiety, depression, autism and learning disability. [22, 25, 54]. As healthcare services recover from the COVID pandemic, future studies will need to determine if these concerning trends in antipsychotic management continue to worsen, to further inform future health and social care policy.

However, we have shown worrying findings: fewer patients are receiving annual psychiatric review, as more are discharged to general practice for sole management. We estimate that in 2020, based on a UK mid-point population estimate of those >18 years of 52 million [58], an antipsychotic prevalence of 1.45%, and 48% of these patients solely cared for by GPs, approximately 360,000 patients are prescribed antipsychotics long-term without psychiatric review. Furthermore antipsychotics are being prescribed for non-psychotic illnesses that are not captured on the QOF SMI register, meaning cardiometabolic monitoring may not occur. There is therefore an urgent need for targeting of resources to enable patients taking antipsychotics long-term to have access to an annual review with a psychiatrist able to optimise or withdraw antipsychotics. Furthermore, general practice funding for cardiometabolic monitoring should be based on antipsychotic use rather than diagnosis. Changes in policy are needed to prevent patients being ‘trapped’ on antipsychotics long-term without psychiatrist review, and ensure adequate cardiometabolic monitoring if we are to address the premature mortality experienced by patients with psychiatric illnesses.

## Ethical Approval

The use of data from the SAIL databank was approved by the SAIL Information Governance Research Panel. As this study did not use person identifiable data no formal NHS research ethics committee approval was required. No data were reported where an output would have <5 individual counts.

## Data Availability

All data produced in the present study are available upon reasonable request to the authors

https://github.com/alanwoodall/AMP-Epidemiology

## Acknowledgements

This study makes use of anonymised data held in the Secure Anonymised Information Linkage (SAIL) Databank. We would like to acknowledge all the data providers who make anonymised data available for research. We want to thank members of SAIL, especially Dr Sarah Rees, who supported the researchers during this project. We are also grateful to consultant psychiatrist colleagues Dr Ben Shooter, and Dr Adnan Sharaf, who provided insight into psychiatric antipsychotic use in the UK, clinic appointment structures, and outpatient recall methods.

## Dedication

This work is dedicated to the memory of our friend and research colleague, Huw Collins, who sadly died unexpectedly during this project.

## Author Information

Alan Woodall, PhD MRCGP, Honorary Clinical Senior Lecturer^1^ and Clinical Lead for Integrated Care^2^, ORCID: 0000-0003-2933-0508;

Alex Gampel, PhD, Information Reporting and Analytics Manager^2^, ORCID: 0000-0001-6923-5818;

Huw Collins, BA, Data Analyst^3^ (deceased; no ORCID/email)

Lauren Walker, PhD MRCP, Reader in Clinical Pharmacology and General Internal Medicine^4^, ORCID: 0000-0002- 3827-4387;

Frances Mair, MD FRCGP FRSE, Norie Millar Professor of General Practice^5^, ORCID: 0000-0001-9780-1135

Sally Sheard, PhD, Andrew Geddes & John Rankin Professor of Modern History, and Wellcome Trust Senior Investigator (Health Policy)^6^, ORCID: 0000-0001-8116-9120;

Pyers Symon, BSc, Public Advisor^7^, ORCID: 0009-0001-6499-8442;

Iain Buchan, MD FFPH, W.H Duncan Professor of Public Health Systems^7^and Director^8^, ORCID: 0000-0003-3392- 1650;

## Abbreviations

APM: Antipsychotic Medication
GP: General Practitioner (Family Medicine physician outside the UK)
NICE: National Institute of Health and Care Excellence
OPDW: Outpatient Dataset for Wales (secondary care outpatient dataset)
PEDW: Patient Episode Dataset for Wales (hospital admission dataset)
QOF: Quality and Outcomes Framework
SAIL: Secure Anonymised Information Linkage
SMI: Serious Mental Illness
WIMD: Welsh Index of Multiple Deprivation
WLGP: Welsh Longitudinal General Practice (dataset)

---

**Table S1:** QOF register contractual requirements in the Primary Care GP Contract. These contractual requirements varied from UK nation to nation over the study period. The table shows the typical monitoring that a patient with a diagnosis would experience according to each register.

|  | QOF Register (annual contract requirements) |  |  |  |
| --- | --- | --- | --- | --- |
|  | SMI | Dementia | Depression | No register |
| **Patients taking antipsychotic with typical diagnoses** | Schizophrenia;<br>Bipolar disorder;<br>Psychotic depression | Vascular dementia;<br>Alzheimer's disease | Moderate depressive disorder | Generalised anxiety disorder;<br>Autism |
| **Registration** | Yes | Yes | Yes | No |
| **GP review** | Yes | Yes | Yes | No |
| **Blood pressure** | Yes | No | No | No |
| **BMI** | Yes | No | No | No |
| **Lipids** | Yes | No | No | No |
| **HBA1c /glucose** | Yes | Yes | No | No |
| **Alcohol consumption** | Yes | No | No | No |

**Table S2:** Counts of patients (all ages) on long-term antipsychotics and licenced indications in UK national formulary.

| Antipsychotic | Read code (5-digit) | Count (2011) | Relative proportion (%) (2011) | Count (2020) | Relative Proportion (%) (2020) | First or Second Generation | Licensed indication in UK British National Formulary (2024) |  |  |  |  |  |  |
| --- | --- | --- | --- | --- | --- | --- | --- | --- | --- | --- | --- | --- | --- |
|  |  |  |  |  |  |  | Psychosis/agitation in Serious Mental Illness (Schizophrenia, Bipolar Disorder, Psychotic Depression) | Psychosis in Parkinson's Disease | Management of Behavioural and Psychological Symptoms in Dementia | Depression (adjunctive treatment, psychosis not required) | Anxiety | Control of deviant antisocial sexual behaviour | Non-psychiatric uses (e.g. nausea) |
| Quetiapine | d4s.. | 6362 | 28.0 | 12338 | 37.1 | 2 | YES | NO | NO | YES | NO | NO | NO |
| Olanzapine | d4r.. OR d58.. | 5430 | 23.9 | 7527 | 22.6 | 2 | YES | NO | NO | NO | NO | NO | NO |
| Risperidone | d4p.. | 3377 | 14.8 | 5106 | 15.3 | 2 | YES | NO | NO | NO | NO | NO | NO |
| Amisulpride | d4t.. | 1531 | 6.7 | 1190 | 3.6 | 2 | YES | NO | NO | NO | NO | NO | NO |
| Aripiprazole | d4v.. | 1381 | 6.1 | 4303 | 12.9 | 2 | YES | NO | NO | NO | NO | NO | NO |
| Haloperidol | d47.. OR d55.. | 997 | 4.4 | 825 | 2.5 | 1 | YES | NO | YES | NO | NO | NO | YES |
| Chlorpromazine | d41.. | 971 | 4.3 | 633 | 1.9 | 1 | YES | NO | NO | NO | YES | NO | YES |
| Trifluoperazine | d4h.. | 669 | 2.9 | 117 | 0.4 | 1 | YES | NO | NO | NO | YES | NO | YES |
| Pericyazine | d4a.. | 576 | 2.5 | 176 | 0.5 | 1 | YES | NO | NO | NO | YES | NO | NO |
| Promazine | d4e.. | 508 | 2.2 | 244 | 0.7 | 1 | YES | NO | NO | NO | NO | NO | YES |
| Sulpride | d4f.. | 490 | 2.2 | 271 | 0.8 | 1 | YES | NO | NO | NO | NO | NO | NO |
| Zuclopenthixol | d4j.. OR d57 | 211 | 0.9 | 257 | 0.8 | 1 | YES | NO | NO | NO | NO | NO | NO |
| Flupentixol | d51.. | 159 | 0.7 | 104 | 0.3 | 1 | YES | NO | NO | YES | NO | NO | NO |
| Benperidol | d42.. | 39 | 0.2 | 34 | 0.1 | 1 | NO | NO | NO | NO | NO | YES | NO |
| Pimozide | d4c.. | 23 | 0.1 | 10 | 0.0 | 1 | YES | NO | NO | NO | NO | NO | NO |
| Clozapine | d4l.. | 14 | 0.1 | 8 | 0.0 | 2 | YES | YES | NO | NO | NO | NO | NO |
| Paliperidone | d4w.. | <5 | 0.0 | 14 | 0.0 | 2 | YES | NO | NO | NO | NO | NO | NO |
| Asenapine | d4x.. | <5 | 0.0 | <5 | 0.0 | 2 | YES | NO | NO | NO | NO | NO | NO |
| Cariprazine | not listed in d4.. | <5 | 0.0 | <5 | 0.0 | 2 | YES | NO | NO | NO | NO | NO | NO |
| Lurasidone | d4y.. | <5 | 0.0 | <5 | 0.0 | 2 | YES | NO | NO | NO | NO | NO | NO |
| Droperidol | d44.. | <5 | 0.0 | <5 | 0.0 | 1 | NO | NO | NO | NO | NO | NO | YES |
| Loxapine | d4k.. | <5 | 0.0 | <5 | 0.0 | 1 | YES | NO | NO | NO | NO | NO | NO |
| Zuclopenthixol acetate | d4n.. | <5 | 0.0 | <5 | 0.0 | 1 | YES | NO | NO | NO | NO | NO | NO |
| Prochlorperazine as antipsychotic (excluded from 'total count as not used as an antipsychotic in UK')* | D4d.. (Dhe.. Antiemetic code use is not captured under d4 code) | <5 | N/A | <5 | N/A | N/A | YES | NO | NO | NO | YES | NO | YES |
| Levomepromazine (excluded from 'total count as not used as an antipsychotic in UK')* | d48.. | 68 | N/A | 158 | N/A | N/A | YES | NO | NO | NO | NO | NO | YES |
| Total (individual antipsychotic counts) |  | 22773 | 100.1 | 33187 | 99.7 | N/A |  |  |  |  |  |  |  |
| Total (top 6 antipsychotic used in 2020) |  | 19078 | 83.9 | 31289 | 94.0 | N/A |  |  |  |  |  |  |  |
| Total 1<sup>st</sup> Generation antipsychotic |  | 4658 | 20.5 | 2686 | 8.1 | N/A |  |  |  |  |  |  |  |
| Total 2<sup>nd</sup> Generation antipsychotic |  | 18115 | 79.7 | 30501 | 91.7 | N/A |  |  |  |  |  |  |  |
| "ALL antipsychotic Total" | codes d4... and d5... excluding any prochlorperazine antiemetic codes (dhe..) and levomepromazine codes | 22119 | N/A | 32086 | N/A | N/A |  |  |  |  |  |  |  |
| All antipsychotics (total as a % of summation of individual antipsychotic count) |  | 97.1 | N/A | 96.7 | N/A | N/A |  |  |  |  |  |  |  |

**Figure S1:**
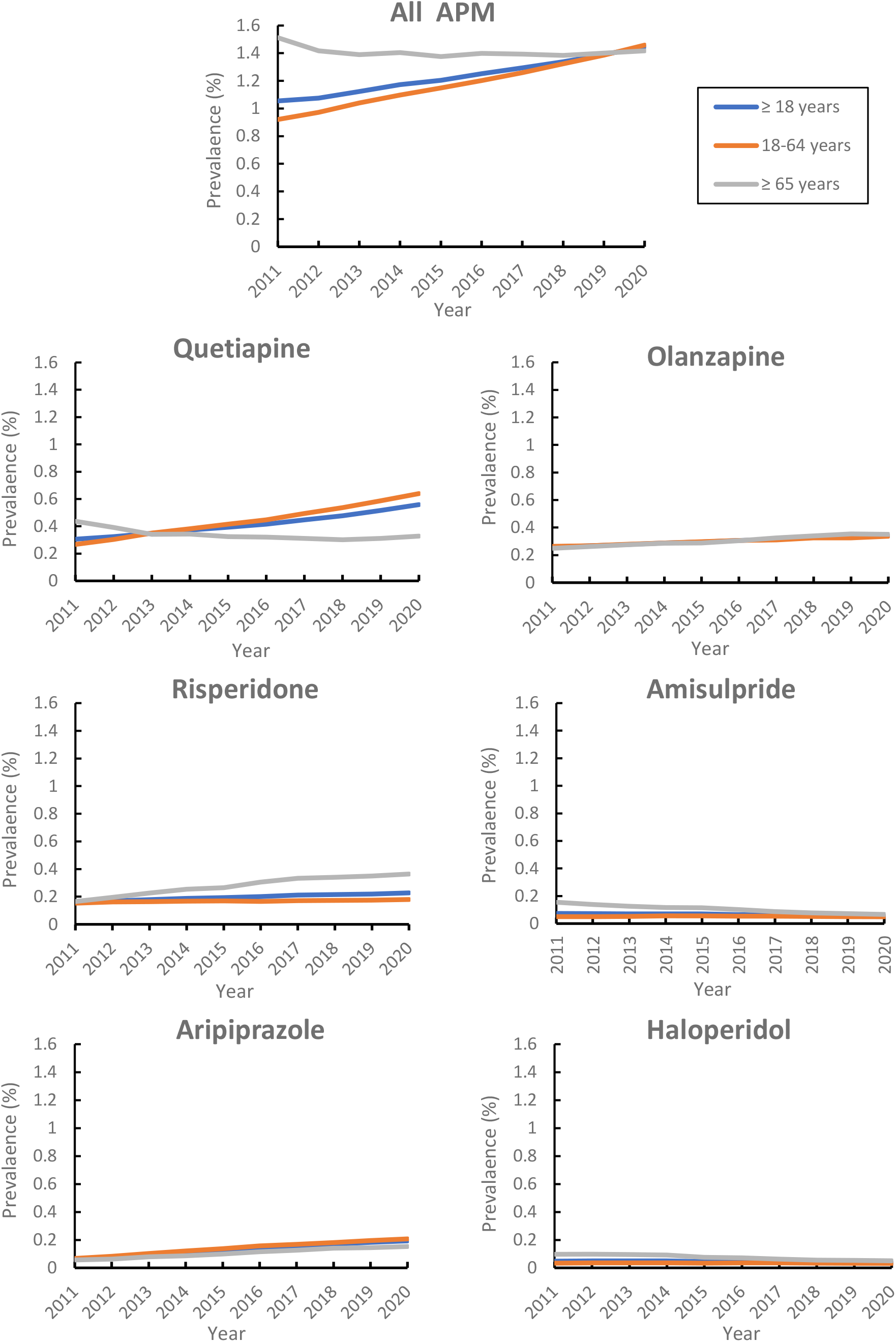
Prevalence of long-term antipsychotic use between 2011 and 2020

**Figure S2:**
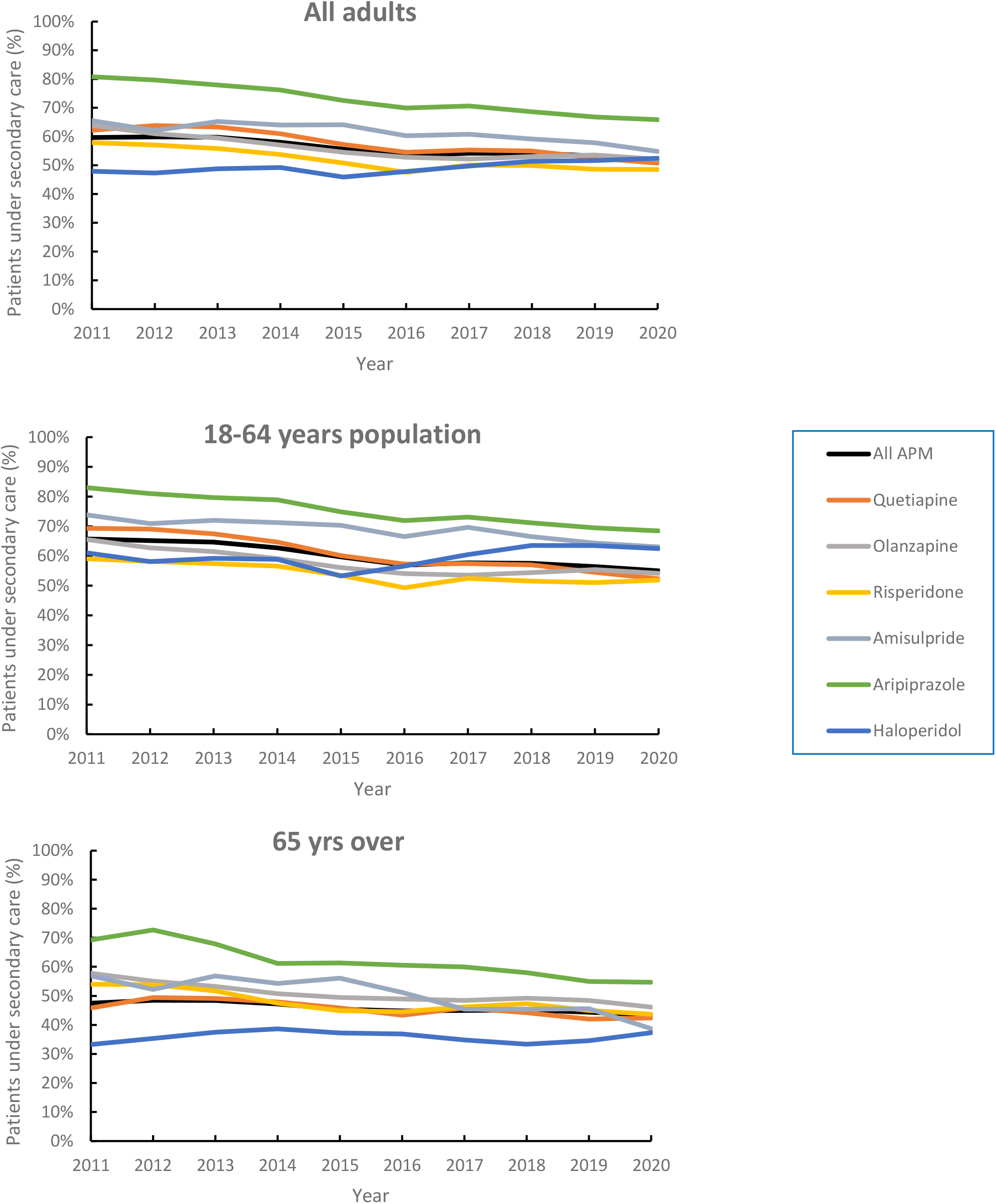
Proportions of patients taking long-term antipsychotics who have had psychiatric review in previous 12 months

**Figure S3:**
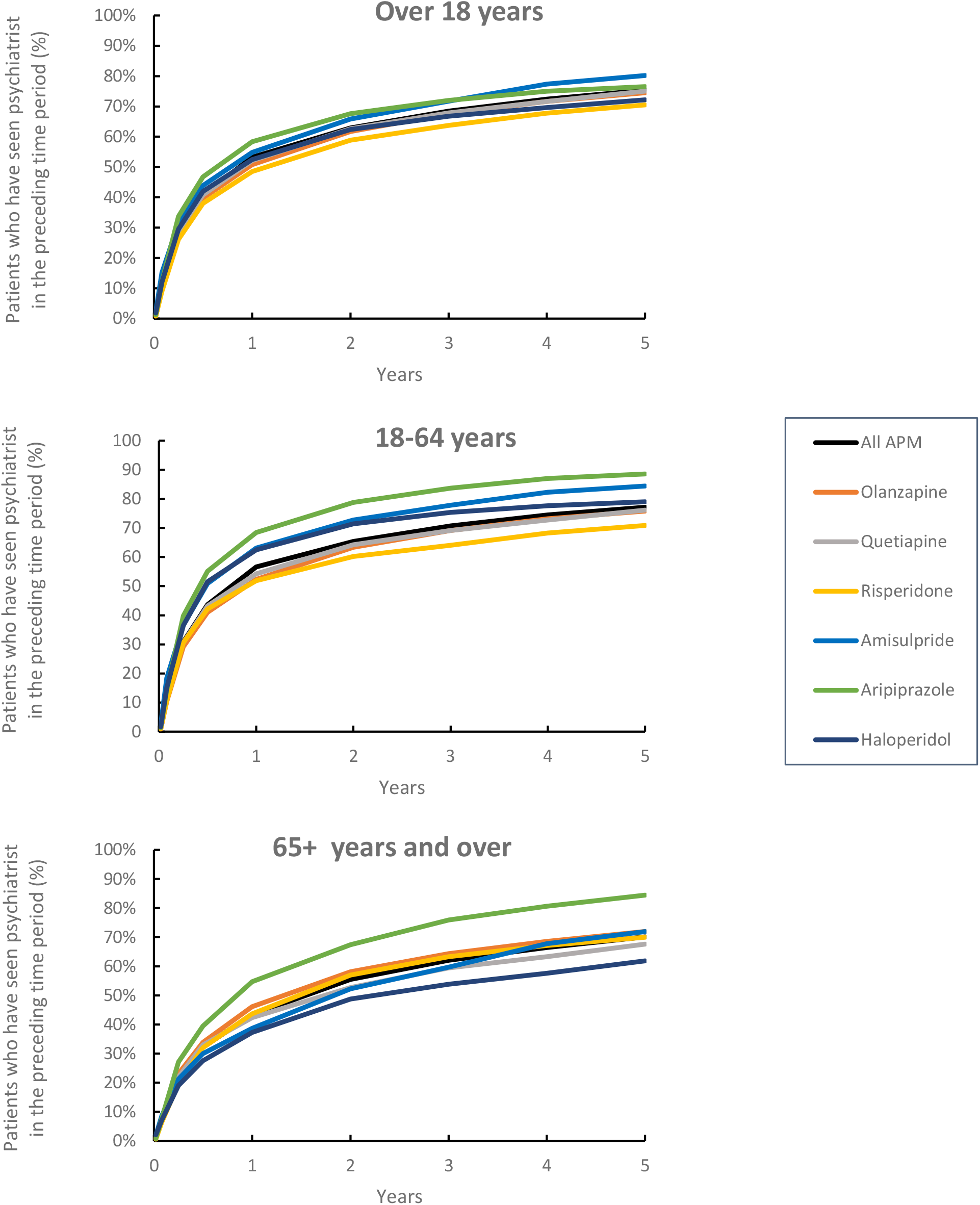
Proportions of patients in 2020 prescribed long-term antipsychotics who have had psychiatric review in the preceding time period of up to 5 years

**Figure S4:**
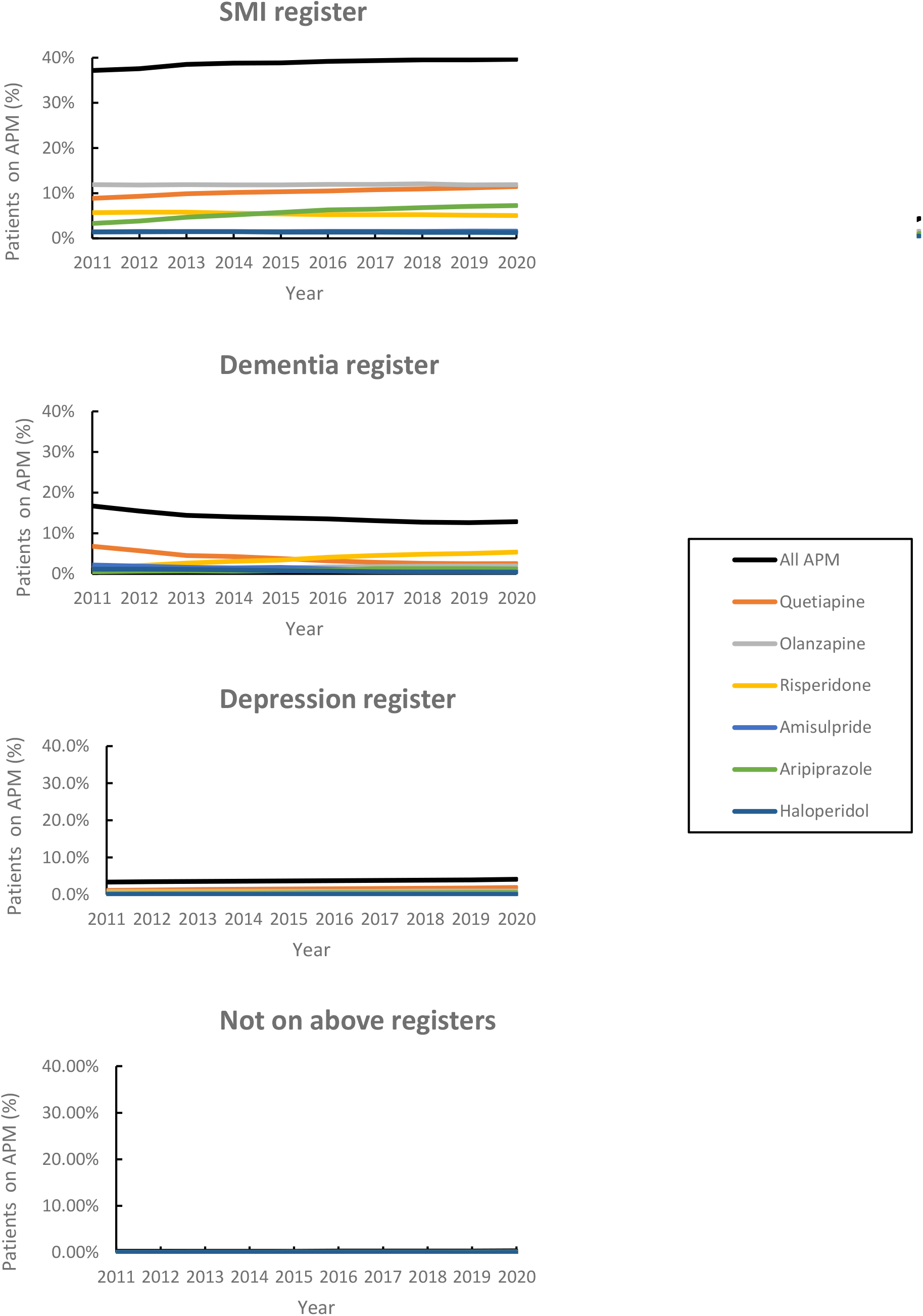
Proportions of patients on QOF registers on long-term antipsychotics

**Figure S5:**
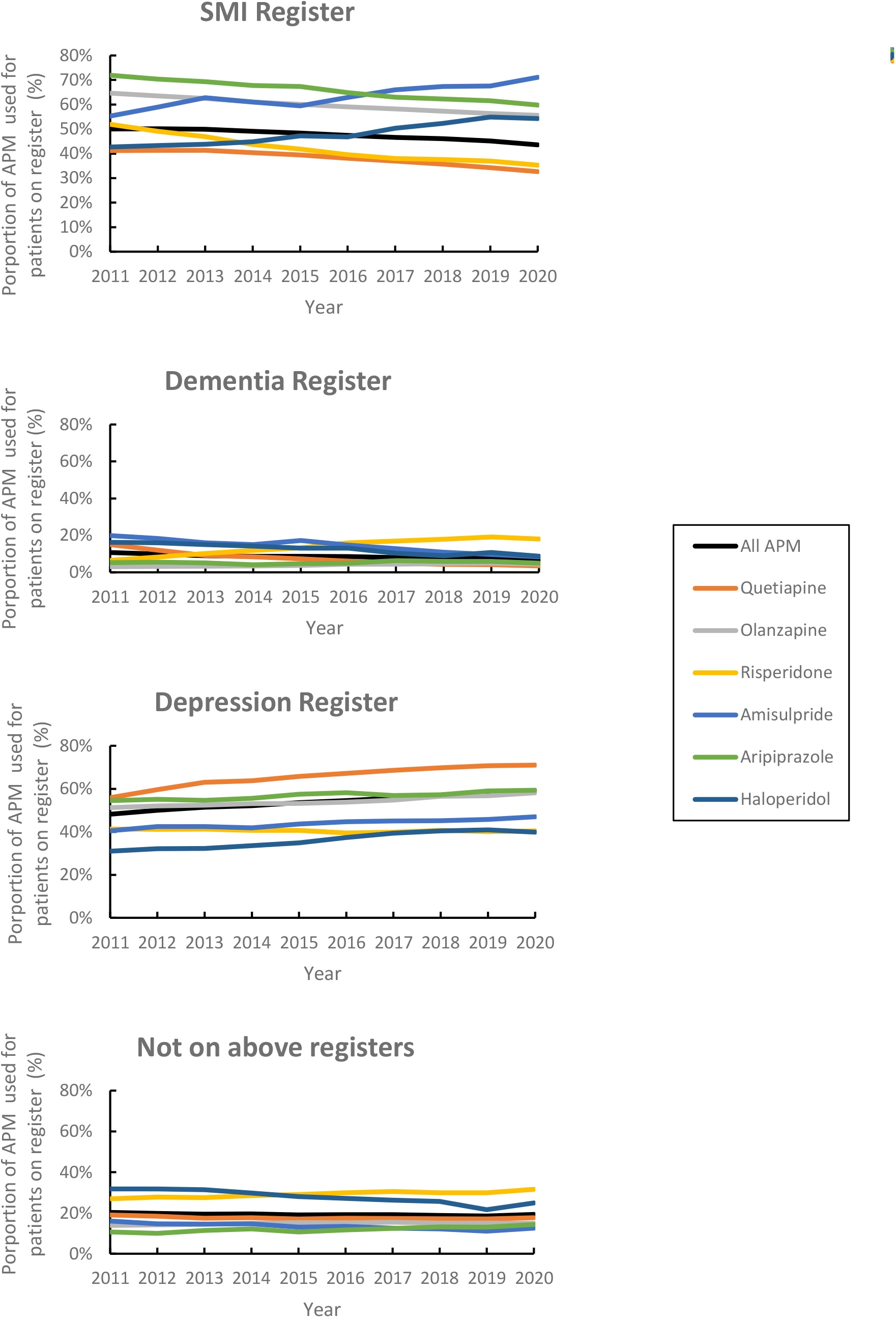
Proportion of overall antipsychotic use prescribed to patients on each QOF register

